# Paratype: A genotyping framework and an open-source tool for *Salmonella* Paratyphi A

**DOI:** 10.1101/2021.11.13.21266165

**Authors:** Arif M. Tanmoy, Yogesh Hooda, Mohammad S. I. Sajib, Kesia E. da Silva, Junaid Iqbal, Farah N. Qamar, Stephen P. Luby, Gordon Dougan, Zoe A. Dyson, Stephen Baker, Denise O. Garrett, Jason R. Andrews, Samir K. Saha, Senjuti Saha

**Affiliations:** Child Health Research Foundation, Dhaka, Bangladesh; Department of Medical Microbiology and Infectious Diseases, Erasmus University Medical Center, Rotterdam, the Netherlands; MRC-Laboratory Molecular Biology, Cambridge, UK; Institute of Biodiversity, Animal Health and Comparative Medicine, University of Glasgow, Glasgow, UK; Division of Infectious Diseases and Geographic Medicine, Stanford University School of Medicine, Stanford, California, USA; Department of Paediatrics and Child Health, Aga Khan University, Karachi, Pakistan; Wellcome Trust Sanger Institute, Hinxton, Cambridge CB10 1SA, UK; Cambridge Institute of Therapeutic Immunology and Infectious Disease, Department of Medicine, University of Cambridge, Cambridge, United Kingdom; Department of Infection Biology, London School of Hygiene and Tropical Medicine, London, UK; Applied Epidemiology Team, Sabin Vaccine Institute, Washington, DC, USA; Department of Microbiology, Bangladesh Institute of Child Health, Dhaka Shishu Hospital, Dhaka, Bangladesh; Department of Infectious Diseases, Central Clinical School, Monash University, Melbourne, Victoria 3004, Australia

**Keywords:** *Salmonella* Paratyphi A, Paratyphoid fever, Paratyphi A genotyping, Genomics, Antimicrobial resistance, Global analysis, Epidemiology, Enteric fever, Neglected tropical disease

## Abstract

**Background:** *Salmonella enterica* serovar Paratyphi A (*Salmonella* Paratyphi A) is the primary causative agent of paratyphoid fever, which is responsible for an estimated 3.4 million infections annually. However, little genomic information is available on population structure, antimicrobial resistance (AMR), and spatiotemporal distribution of the pathogen. With rising antimicrobial resistance and no licensed vaccines, genomic surveillance is important to track the evolution of this pathogen and monitor transmission.

**Results:** We performed whole-genome sequencing of 817 *Salmonella* Paratyphi A isolates collected from Bangladesh, Nepal, and Pakistan and added publicly available 562 genomes to build a global database representing 37 countries, covering 1917-2019. To track the evolution of *Salmonella* Paratyphi A, we used the existing lineage scheme, developed earlier based on a small dataset, but certain sub-lineages were not homologous, and many isolates could not be assigned a lineage. Therefore, we developed a single nucleotide polymorphism based genotyping scheme, Paratype, a tool that segregates *Salmonella* Paratyphi A into three primary and nine secondary clades, and 18 genotypes. Each genotype has been assigned a unique allele definition located on a conserved gene. Using Paratype, we identified genomic variation between different sampling locations and specific AMR markers, and mutations in the O2-polysaccharide synthesis locus, a candidate for vaccine development.

**Conclusions:** This large-scale global analysis proposes the first genotyping tool for *Salmonella* Paratyphi A. Paratype has already been released (https://github.com/CHRF-Genomics/Paratype) as an open-access, command-line tool and is being adopted for large scale genomic analysis. This tool will assist future genomic surveillance and help inform prevention and treatment strategies.

## Background

Paratyphoid fever, caused by *Salmonella enterica* subspecies *enterica* serovar Paratyphi A (*Salmonella* Paratyphi A) is a systemic febrile illness that affects an estimated 3.4 million people each year, and causes 19,100 deaths globally [1]. The disease is clinically indistinguishable from typhoid fever, caused by *Salmonella enterica* subspecies *enterica* serovar Typhi (*Salmonella* Typhi). Much like typhoid, paratyphoid fever is also endemic in many low- and middle-income countries of South Asia and Sub-Saharan Africa, due to fecal contamination of water, food and the environment. However, barring a few countries (e.g., China, Myanmar), paratyphoid fever is usually less prevalent than typhoid fever [2,3]. *Salmonella* Paratyphi A continues to be an inadequately studied pathogen [4] hampering the implementation of evidence-based policies for the treatment and prevention of paratyphoid fever.

Relative to *Salmonella* Typhi, little genomic information is available on population structure, antimicrobial resistance (AMR), and spatiotemporal distribution of *Salmonella* Paratyphi A. The first *Salmonella* Paratyphi A genome was published in 2004 and had a size of 4.5 Mb, with ∼4,200 genes. To determine the global diversity of *Salmonella* Paratyphi A isolates, Bayesian analysis was conducted on a set of 149 *Salmonella* Paratyphi A genomes, which identified that the last common ancestor of all *Salmonella* Paratyphi A existed for at least 450 years prior to differentiating into at least seven distinct lineages (A to G) which have circulated globally [5]. Whole genome sequencing was also used to characterize clonal paratyphoid outbreaks in Cambodia [6] and China [7] and further extend the lineage scheme to include sub-lineages within Lineage A and C. However, very few studies have characterized isolates from countries in South Asia, which contributes over 80% of all paratyphoid infections [8,9]. Available studies are sporadic, and either focused on genomes from a specific geographical location or provide no information on antimicrobial resistance markers, potential vaccine targets, and other virulence factors.

To address this data gap, we performed whole-genome sequencing of 817 *Salmonella* Paratyphi A isolates collected from Bangladesh, Nepal, and Pakistan and combined them with whole-genome sequence data of another 562 isolates reported in the literature to build a global database of 1,379 *Salmonella* Paratyphi A isolates. To track the evolution of *Salmonella* Paratyphi A over a century, we used the existing lineage scheme and found that certain sub-lineages were not homologous, and many isolates could not be assigned a specific lineage. This motivated us to develop a single nucleotide polymorphism (SNP) based genotyping scheme, called Paratype. The scheme is phylogenetically informative and successfully segregates the global population structure into three primary, seven secondary, and 18 distinct subclades/genotypes. We also identified the specific antimicrobial resistance genes, mutations, and plasmids present in *Salmonella* Paratyphi A genomes and correlated these with the different genotypes.

## Results

### Whole-genome sequencing and compilation of global *Salmonella* Paratyphi A genomes

The Child Health Research Foundation (CHRF) has been conducting typhoid and paratyphoid fever surveillance in Bangladesh since 1999 and has generated a biobank of 1,123 *Salmonella* Paratyphi A isolates from 1999-2018 [10–12]. We selected 528 of these isolates, covering all age groups, years of isolation, and hospitalization status (hospitalized/out-patient), and performed whole-genome sequencing on these isolates (Additional file 1: Table S1). Of these, 180 S*almonella* Paratyphi A isolates were collected as part of the Surveillance of Enteric Fever in Asia Project (SEAP, 2014 - 2019) study, a multi-country international effort to better understand the epidemiology and impact of enteric fever in South Asia [13]. In addition to Bangladesh, 133 isolates were sequenced from the SEAP study conducted in Pakistan, and 156 from Nepal.

To contextualize these genomes, we conducted a literature search to compile all publicly available *Salmonella* Paratyphi A genomes (for which raw reads were available) to build a database of 560 additional isolates from 10 studies (Additional file 1: Table S2). Two reference genomes (ATCC 9150 and AKU_12601) were also included. The largest dataset consisted of 254 isolates, published by Public Health England as part of their *Salmonella* surveillance [8,14]; 164 of these isolates were linked to travel, most commonly to South Asia. In our study, we assigned these isolates to the countries where the patient acquired the infection. Our final data, including the genomes we sequenced, consisted of a total of 1,379 isolates from 37 different countries, spanning over 103 years - 1917 to 2019. Most of the isolates (1,112/ 1,379; 81%) were from countries in South Asia (541 from Bangladesh, 268 from Nepal, 187 from Pakistan and 115 from India). South Asian countries also bear a disproportionately high burden of paratyphoid fever; of the estimated 3.4 million global paratyphoid infections in 2019, 2.8 (82%) million are estimated to have occurred in South Asia [1].

Following assembly from raw reads, the pan-genome analysis identified 6,983 genes, of which 4,114 (59% of all genes) were conserved in more than 95% of isolates (Additional file 1: Figure S1). The average genome size was 4.5 Mb with ∼4,300 genes, and the pan genome does not appear to be closed (decay parameter, alpha = 0.67). Overall, 2,550 genes were found to be present in less than 15% of isolates, and these included genes often found in prophages and other mobile regions, and genes encoding adhesins, antimicrobial resistance markers, and several hypothetical proteins.

### Genotyping scheme for *Salmonella* Paratyphi A

To investigate the genomic diversity of *Salmonella* Paratyphi A, we identified 8,346 single nucleotide polymorphisms (SNPs) in the 1,379 isolates. These were used in RAxML [15] to generate a Maximum-likelihood phylogenetic tree of the global collection of *Salmonella* Paratyphi A isolates (Figure 1). A previously reported lineage scheme, proposed for *Salmonella* Paratyphi A by Zhou et al. [5] and extended by subsequent studies [6,7,9,16,17] was overlaid on the RAxML tree. This highlighted the insufficiency of the current lineage scheme to fully capture the diversity of *Salmonella* Paratyphi A present. First, while the isolates from lineages B & D - G clustered together, several isolates previously assigned to lineages A and C in the scheme did not. Second, some sequences belonged to clades that diverged from isolates before the exitance of the most recent common ancestor for lineages A and B, indicating that these isolates should be considered to be in a different lineage. This was not surprising considering that when this scheme was devised, there were limited number of sequenced *Salmonella* Paratyphi A genomes available, particularly from South Asia.

**Figure 1:**
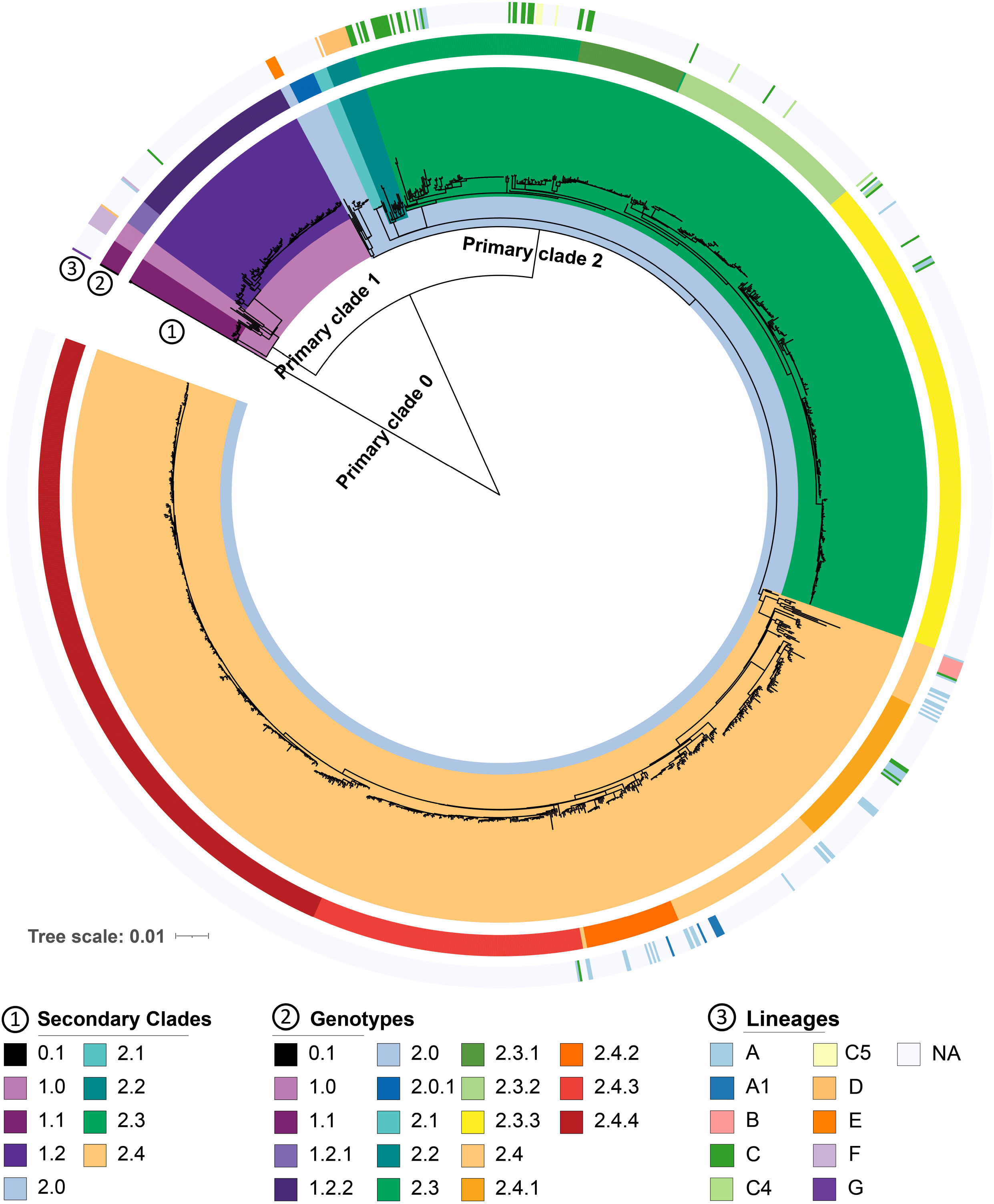
Genotyping scheme for *Salmonella* Paratyphi A. The scheme is composed of three primary, nine secondary and 18 genotypes on a phylogenetic tree of 1,379 isolates. The 9 secondary clades as highlighted by the coloring of the inner ring. 18 genotypes identified and are shown in the colored middle ring of the figure. The previously proposed lineage system is shown in the outer ring.

To build a genotyping scheme based on a larger number of representative samples, first, we used fastBAPS [18] to generate a potential list of clusters in the RAxML tree (Additional file 1: Figure S2). Next, we selected a set of 315 isolates that included two isolates per year for all fastBAPS clusters selected randomly and performed phylodynamic analysis using the Bayesian Evolutionary Analysis by Sampling Trees (BEAST) software [19] (Figure 2). Based on these analyses, we devised a genotyping scheme with three primary clades, nine secondary clades, and 18 genotypes that have circulated globally in the last 100 years.

**Figure 2:**
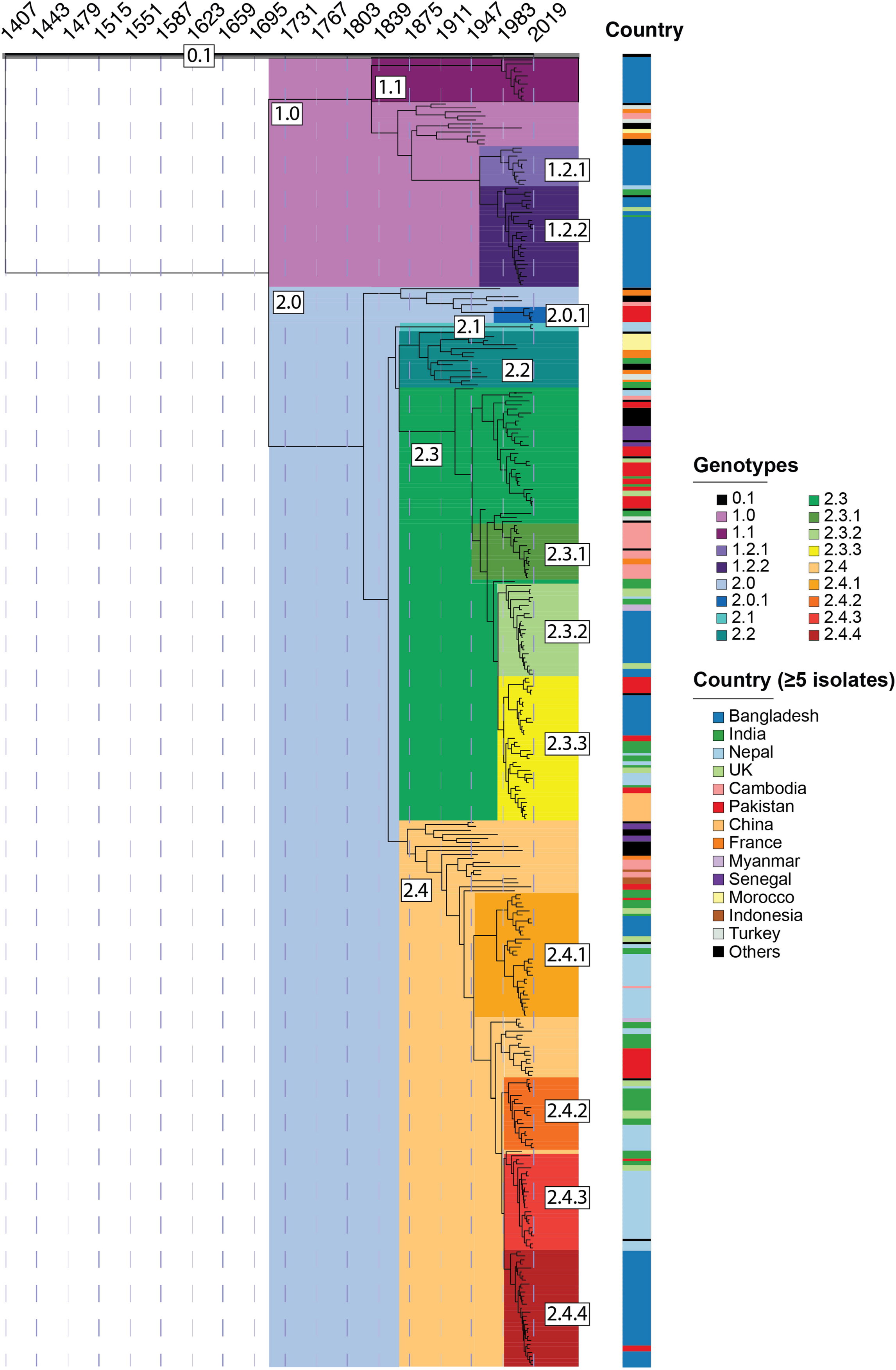
Maximum clade credibility tree of 315 representative *Salmonella* Paratyphi A isolates. The tree shows the last common ancestor of all *Salmonella* Paratyphi A existed at least 600 years ago (tMRCA - 1407 AD). The different genotypes are temporally resolved. Countries with greater than or equal to 5 isolates are also included.

To aid further genomic epidemiological studies, we identified 18 additional alleles (Additional file 1: Table S3) that are unique to each of the 18 *Salmonella* Paratyphi A genotypes. These alleles were present in conserved genes involved in essential cellular functions such as protein synthesis, DNA replication, or metabolism. Identification of these genotype-specific alleles allowed us to write a Python script – “Paratype” – that assigns genotypes to *Salmonella* Paratyphi A genomes using fastq, bam, or vcf files obtained during whole-genome sequencing and variant calling. The Paratype software tool (available at: https://github.com/CHRF-Genomics/Paratype/) has 100% sensitivity and specificity and was able to assign the correct genotype to all the 1,379 genomes that were present in our database.

### Temporal and geographic distribution of different genotypes

Upon the establishment of the “Paratype” scheme, we considered the geographical distribution of the different genotypes (Figure 3). Genotype 0.1 under primary clade 0 was phylogenetically unique (matches with lineage H of Zhou et al [5]); there was only one isolate belonging to this genotype/primary clade that was isolated in Hong Kong in 1971. The genome of this isolate was distinct from all other genomes obtained thus far, contained 1288 unique SNPs, and may represent a lineage that is now extinct, or present at very low numbers in areas that have not been sampled. The other two primary clades, clades 1 and 2, emerged between 1700-1800 and contain genomes that have been collected in the last two decades. Clade 1 contains strains largely from lineage F, and fastBAPS predicted two sub-clusters within this clade. One of these clusters was largely found in Bangladesh and has been assigned secondary clade 1.2, then sub-divided into genotypes 1.2.1 and 1.2.2 which appear to have diverged in the 1950s. Both these genotypes are currently present in Bangladesh and other South Asian countries (Figure 2). The other cluster with13 genomes from Bangladesh that were first isolated in 1999 have been assigned to genotype 1.1. The remaining 10 genomes were obtained between 1917 to 1963 and have been assigned genotype 1.0.

**Figure 3:**
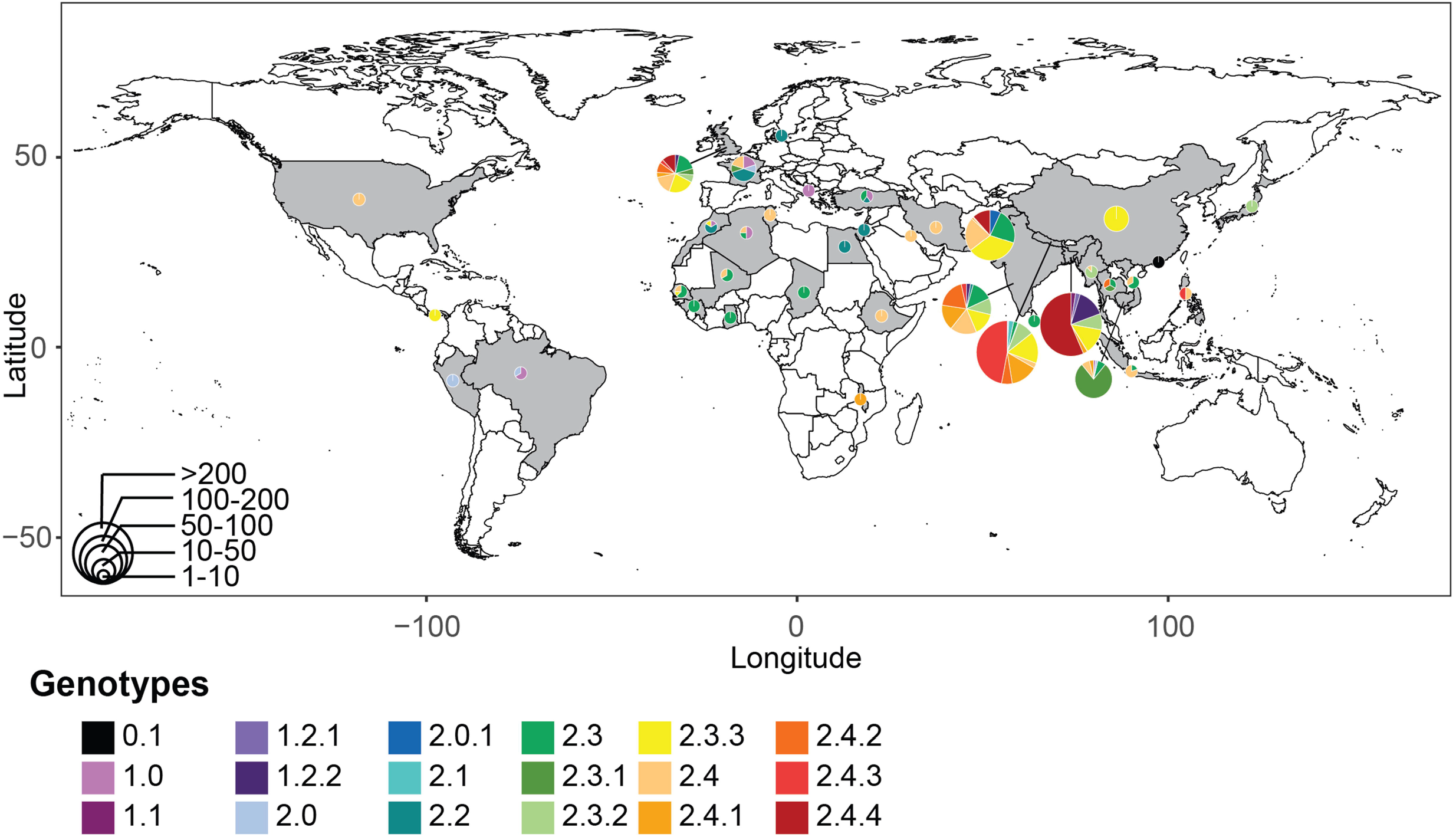
Geographical distribution of *Salmonella* Paratyphi A genotypes. The country of isolation for 1378 sequenced *Salmonella* Paratyphi A isolates is shown. The distribution of genotypes per country is shown as scattered pie charts. The size of the each pie chart represents the number of sequences available. A difference in circulating genotypes is observed indicating local populations differ in several endemic countries. Further details are provided in Additional file 2.

Most *Salmonella* Paratyphi A genomes (1254/1379; 91%) have been assigned to primary clade 2, which contains genomes belonging to the lineage A-E of the previous scheme. Genomes that belonged to lineages B, D, and E have now been assigned to genotypes 2.4, 2.2, and 2.0, respectively. Within genotype 2.0, 13 unique and recent isolates from Pakistan were identified and have been assigned as genotype 2.0.1. Genotype 2.1 contains isolates from Nepal that were sampled during the SEAP study, yet the genotype emerged in the 1800s and is distinct from all other isolates in clade 2. Two clusters in fastBAPS, comprising of strains largely from what was formerly C lineage are now assigned to genotype 2.3. Genotype 2.3 has been subdivided into genotypes 2.3.1 to 2.3.3, each of which belongs to a distinct geographical location: 2.3.1 is found predominantly in Cambodia and South-East Asia; 2.3.2 and 2.3.3 are found largely in South Asia. An outbreak of paratyphoid fever in China during 2010 – 2011 [7] was caused by isolates of genotype 2.3.3, and these likely originated in South Asia. The former lineages A and B have been assigned genotype 2.4, which is further divided into 2.4.1 to 2.4.4. While genotypes 2.4.1 and 2.4.2 have been observed in different countries in South Asia, genotype 2.4.4 is predominantly found in Bangladesh, and 2.4.3 is largely present in Nepal.

Different countries in South Asia had unique genotype distributions. Predominant genotypes present in Bangladesh were 2.4.4 (56%) followed by 1.2.2 (14%) and 2.3.3 (13%). In Nepal, 2.4.3.(47%), 2.3.3 (16%) and 2.4.1 (14%) were three most common genotypes. Pakistan had genotypes 2.3.3 (25%), 2.3 (16%) and 2.4 (15%). In India, genotypes 2.4.2 (22%), 2.4 (20%), 2.4.1 (19%), 2.3.3 (17%), and 2.3 (16%) were commonly identified.

### Antimicrobial resistance markers in *Salmonella* Paratyphi A

To characterize genomic determinants of antimicrobial resistance in *Salmonella* Paratyphi A, we screened the 1,379 genomes for the presence of antimicrobial genes and markers using ResFinder [20] (Figure 4a) and plasmids using PlasmidFinder [21] (Figure 4b). Of the 1,379 isolates, 1,015 (74%) isolates showed no predicted plasmids and 1356/1379 had no predicted antimicrobial resistance genes. Five genomes with the IncHI1 plasmid were identified, two genomes (both from India) contained resistance genes for trimethoprim and chloramphenicol, and the other three genomes contained genes for trimethoprim, chloramphenicol and ampicillin designated as MDR isolates (one each from India, Pakistan, and Thailand). All five genomes belonged to genotype 2.3 and the strains were isolated between 1999-2004. We also identified a genome belonging to genotype 2.4.4 containing *bla*CTX-M-15 and *bla*TEM-1B on an IncI1-I plasmid; the originating strain was isolated from a patient who contracted the infection in Bangladesh in 2017 [22]. There were 14 isolates from the genotype 2.3.1 that contain *bla*TEM-116, which can lead to resistance to ampicillin; all 14 were reported from Cambodia[6]. Another isolate from genotype 2.3.3 (from Pakistan, 2015) contained a *qnrB19* gene on a Col(pHAD28) plasmid, which has been shown to lead to quinolone resistance in other *Salmonella* species [23].

**Figure 4:**
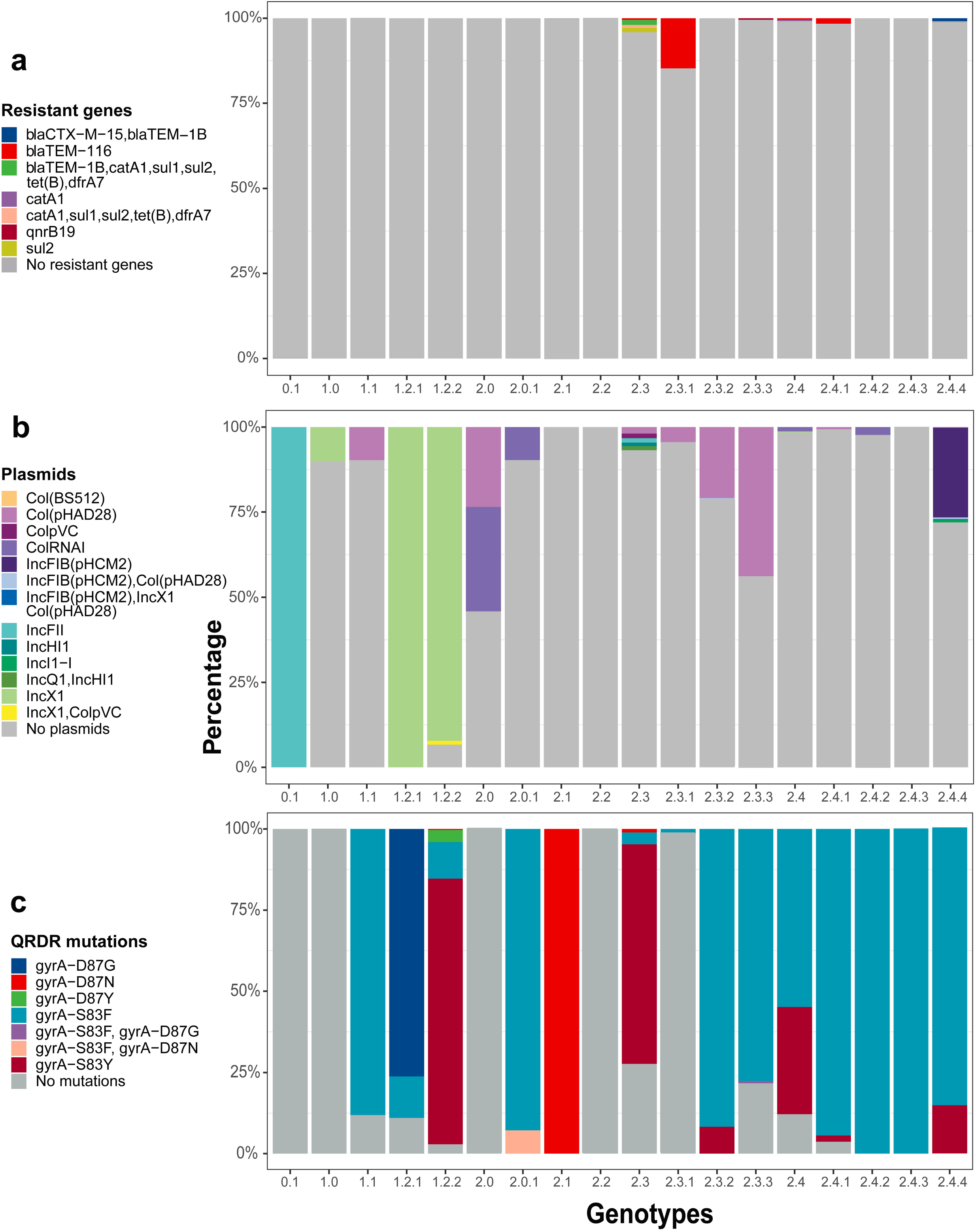
Presence of antimicrobial resistance genes, plasmids, and chromosomal mutations linked to quinolone resistance across different *Salmonella* Paratyphi A genotypes. The diversity of **a)** Antimicrobial resistance genes **b)** Plasmids and **c)** quinolone resistance determining region (QRDR) mutations present *Salmonella* Paratyphi A is shown.

In addition to antimicrobial resistance genes, we also identified chromosomal mutations in the *acrB* gene and the quinolone resistance determining region (QRDR) to identify isolates resistant to azithromycin and ciprofloxacin respectively. Six of 1,379 genomes contained an AcrB R717 mutation, all from Bangladesh and these belonged to genotypes 2.3.3 (1/6) and 2.4.3 (5/6) [24,25]. The first azithromycin resistant *Salmonella* Paratyphi A isolate was identified in 2014, and this resistance has emerged independently at least twice in two different genotypes. On the other hand, a majority (1177/1397; 84%) of genomes had mutations in the QRDR region. The most commonly found single mutation was gyrA-S83F (941/1379), followed by gyrA-S83Y (205/1379). Two isolates contained double mutations in the QRDR region; one of them belonged to genotype 2.0.1 (gyrA-S83F & D87N, Pakistan, 2017) and another belonged to genotype 2.3.3 (gyrA-S83F & D87G, UK, 2016). Barring genotype 0.1, 1.0 and 2.2, all other genotypes had at least one genome with a QRDR mutation (Figure 4c). The first QRDR mutation was identified in 1997 in India in genotype 2.4 and their prevalence have increased over time. In 2012 and 2013, there was an outbreak in Cambodia caused by a strain from genotype 2.3.1 that did not have any QRDR mutation leading to a temporary increase in proportion of *Salmonella* Paratyphi A with no QRDR mutations during these two years (Additional file 1: Figure S3).

### Characterization of mutations in the O2-antigen biosynthetic gene cluster

The majority of the vaccines being developed for *Salmonella* Paratyphi A use the O2-antigen that is unique to this serovar conjugated to a carrier protein [26]. Recently, through in-silico metabolic reconstruction, an 18.9 kb region containing genes involved in O-antigen biosynthesis was identified as important for determining the specific molecular features of the O2-antigen found in *Salmonella* Paratyphi A [27]. We identified the SNPs in the O2-antigen biosynthesis genes found in the 1,379 genomes to investigate the conservation of this genomic loci. In total, 84 SNPs were found, of which 13 were present in more than 10 genomes. The most common SNP was at genomic location 8,68,444 (G> C; synonymous mutation in *prt* gene encoding paratose synthase), which was found in 17% (239/1,379) of all isolates. Out of those 13 common SNPs (n≥10), seven led to non-synonymous mutations (Additional file 1: Figure S4) that could potentially change the O2-antigen structure and chemistry.

## Discussion

*Salmonella* Paratyphi A is the causative agent of paratyphoid fever, a neglected tropical disease with a high burden and mortality in low-and-middle-income countries. Limited information is available regarding its genomic diversity, especially from South Asian countries that collectively are responsible for over 80% of all paratyphoid cases. As genomic surveillance becomes more prominent, there is a need for a coherent and easy-to-use scheme that can be deployed by public health researchers that do not require extensive compute resources.

We sequenced 817 isolates from Bangladesh, Pakistan and Nepal collected over the last 20 years and compiled a collection of all genomes of *Salmonella* Paratyphi A publicly available thus far. We describe a genotyping framework for *Salmonella* Paratyphi A using 1,379 isolates obtained from 1917 through 2019. Rather than being guided by a single approach, we combined ML-based phylogenetics with BAPS and Bayesian analysis via BEAST to design a genotyping scheme for *Salmonella* Paratyphi A. The scheme divided the *Salmonella* Paratyphi A population into 18 different genotypes, and each can be identified by the presence of an allele that is located on the coding sequence of a conserved gene, involved in housekeeping functions. We only found 8,346 SNPs from all 1,379 isolates, with minimal recombination, and thus, this genotyping scheme based on SNP alleles can support robust genotyping and accommodate future evolution of *Salmonella* Paratyphi A. And to assist with that, we have developed Paratype, an open-source Python script for genotyping of *Salmonella* Paratyphi A genomes. Paratype can detect the genotype of *Salmonella* Paratyphi A genomes directly from raw fastq read data. It can also detect mutations in the acrB efflux pump (determinant of macrolide resistance) and in the QRDR region (determinant of ciprofloxacin non-susceptibility).

In this genotyping scheme, we propose three primary clades 0, 1, and 2, which diverged before the 1800s (Figure 2). While only a single isolate of primary clade 0 was obtained in 1971, isolates belonging to clade 1 and 2 have been routinely identified over the past two decades. Clade 2 is the most abundant and has been subdivided into four secondary clades: 2.1 - 2.4, which probably emerged in the 1800s or the early 1900s. Clade 2.3 could be subdivided into 2.3.1 - 2.3.3, each with distinct geographic distribution. Clade 2.4 was also sub-divided into genotypes 2.4.1 - 2.4.4. Genotype 2.4.4 was the most abundant and was predominantly present in Bangladesh. This genotype emerged in the late 1990s to early 2000s and possesses high rates of ciprofloxacin resistance (Figure 2). Five of the isolates from this genotype also contained AcrB-R717Q mutation that leads to azithromycin resistance, while one was found to harbor a plasmid containing extended-spectrum beta-lactamase gene (*bla*CTX-M-15) [22].

In line with findings of previous studies, the rates of acquisition of antimicrobial resistance markers in *Salmonella* Paratyphi A is lower relative to *Salmonella* Typhi (Figure 4) [6,9]. Although a few isolates did acquire the IncHI1 plasmid in the late 1990s to early 2000s (Figure 4a), no massive spread across the globe was noted; this unlike *Salmonella* Typhi lineage H58 (genotype 4.3.1) carrying the IncHI1 plasmid spread and became the dominant lineage in the last 30 years[28]. This is also true for chromosomal mutations such as QRDR and AcrB mutations, which are overall less prevalent in *Salmonella* Paratyphi A than in *Salmonella* Typhi [28,29]. Considering the genetic similarities between *Salmonella* Typhi and Paratyphi A, and the fact that they occupy the same environmental niche, the differences in the presence of AMR genes between these typhoidal *Salmonella* serovars warrants further investigation.

The specific O-antigen in the *Salmonella* Paratyphi A is thought to be conserved (assigned to serogroup O2) and several vaccine candidates are currently under development, utilizing the O2 antigen conjugated to a carrier protein as the main vaccine antigen. We compared the 18.9 kbp region responsible for the synthesis of the O2 antigen in this serovar [27] and found 83 SNPs in this region, of which 7 non-synonymous mutations were present in >10 isolates. While it is not clear if these mutations affect the O2-antigen chemistry, the low mutation rate and no observed recombination events in the cluster suggests that the O2 antigen vaccine will have a broadly protective response against all the *Salmonella* Paratyphi A genotypes sampled thus far. However, any variations in this region should be carefully monitored through genomic surveillance.

The conclusions that we can draw from this analysis are subject to certain limitations. First, the available genomes are an incomplete sample; *Salmonella* Paratyphi A is a neglected pathogen, and hence the available genomes and might not have broad representativeness across geographies or time. Specifically, a small proportion of genomes were available from countries in sub-Saharan Africa and India. Second, while the tool has high sensitivity and specificity on our dataset, as more genomes become available over time and novel mechanism of AMR emerge, this tool will require updates from the bigger scientific community. Like all genotyping tools, Paratype is a living tool that will require updates. Our diverse group of authors plans to continually monitor the library of publicly available genomes, accept update requests via GitHub, and incorporate any required updates in the Paratype scheme accordingly.

## Conclusions

This study reports the first large-scale global analysis of *Salmonella* Paratyphi A genomes and proposes the first genotyping tool for this pathogen. Paratype, which has already been released (https://github.com/CHRF-Genomics/Paratype) as an open-access, easy-to-use, command-line tool, is being tested and adopted by researchers for large scale genomic analysis (https://doi.org/10.5281/zenodo.5520408). This tool will assist future genomic surveillance studies and will help inform prevention and treatment strategies for this neglected pathogen.

## Methods

### Study site and isolate selection

Child Health Research Foundation in Bangladesh has been preserving invasive *Salmonella* isolates since 1999 and maintains a biobank of >9000 typhoidal *Salmonella* isolates, largely from children (<18□years of age) that were isolated from the blood of the patients in two different settings: in-patient (hospitalized), and out-patient (community) facility [30]. Clinical and epidemiological data were collected for all hospitalized patients. From this biobank, among 640 *Salmonella* Paratyphi A isolates collected till December 2016, 348 were randomly selected for whole-genome sequencing (WGS) (Additional file 1: Table S1). A set of 469 *Salmonella* Paratyphi A isolates were also added to this collection, isolated under the Surveillance for Enteric Fever in Asia (SEAP) project from three different typhoid-endemic countries, Bangladesh (n= 180), Nepal (n= 156), and Pakistan (n=133). The SEAP-Bangladesh isolates (n=180) were selected using randomization to represent 483 isolates collected between 2016 and 2018. In contrast, SEAP-Nepal isolates included all pre-SEAP isolates (2014 – 2016) and randomly selected SEAP isolates (2017 – 2019). The SEAP-Pakistan isolates were selected prioritizing the availability of geographic information and susceptibility profile during 2016 – 2018.

To add to all the isolates sequenced in this study, we also collected raw fastq data of 560 *Salmonella* Paratyphi A isolates from 37 different countries and 10 published articles (Additional file 1: Table S2). Complete chromosomal sequences of *Salmonella* Paratyphi A ATCC 9150 (NC_006511) and AKU_12601 (NC_011147) were also included [31,32]. For travel-related paratyphoid cases, the country of “traveling from” was considered as the country of origin. If no travel data is available, the country of “reported from” was considered as the country. Overall, for globally distributed 562 *Salmonella* Paratyphi A, year and country data were available for 507 and 536 respectively (Additional file 1: Table S2). In total, we obtained a global collection of 1,379 *Salmonella* Paratyphi A covering a timeline of 1917 – 2019 and 37 countries [see Additional file 2 for more details].

### Whole-genome sequencing

*Salmonella* Paratyphi A isolates from 1999-2016 (before the start of the SEAP project) from Bangladesh (n=348) were subcultured on MacConkey agar media and kept overnight at 37°C. In case of any visible contamination, a single colony was picked and subcultured again. Later, all colonies were swapped and resuspended into 1 ml of water. From this suspension, 400 µL was used for DNA extraction using the QIAamp DNA Mini Kit (Qiagen, Hilden, Germany) and sent to Novogene (NovogeneAIT, Singapore) for WGS on Novaseq 6000 platform (PE150). All SEAP isolates were extracted using the same protocol and were sequenced on Illumina HiSeq 4000 platform (PE150) at the Welcome Sanger Institute, Cambridge, UK.

### Systematic literature review of existing *Salmonella* Paratyphi A genomes

To contextualize the genomes sequenced in this study, we conducted a systematic search to compile all publicly available *Salmonella* Paratyphi A genomes (for which raw reads and metadata were available) to build a database of 560 additional isolates from 10 studies (Additional file 1: Table S2). First, the search terms “(Salmonella Paratyphi A) AND (Molecular Epidemiology)” “Salmonella Paratyphi A genome” and “(Salmonella Paratyphi A) AND (Genomic Epidemiology)” were used in PubMed advanced search builder. Next, the hits were filtered by selecting dates between 1900 and 2019 and the total number of publications remaining were 231. After screening the abstracts and titles manually and eliminating duplicated, only 7 studies were found to have any kind of genome/metadata available for further analysis. In addition, three studies [8,9,22] that meets our criteria (published and both metadata and raw reads available) but missed/not published during the initial PubMed search were incorporated from European Nucleotide Archive (ENA) database, taking the final number of incorporated publications to 10.

### Quality check, genome assembly, annotation, and pan-genome analysis

Raw fastq reads of all *Salmonella* Paratyphi A were quality-checked using FastQC and trimmed using Trimmomatic if necessary[33]. All 1,377 sets of raw fastq reads were assembled using Unicycler v0.4.8 (*default with --min_fasta_length 200*)[34]. The assembled contigs (n = 1,377) and downloaded complete chromosomes (n = 2) were annotated using Prokka (*--gcode 11 --mincontiglen 200*) [35]. The annotated GFF files of all 1,379 isolates were used to build a pan- and core-genome of *Salmonella* Paratyphi A using Roary v3.3 (*options: -t 11 -e --mafft -n*)[36]. The gene_presence_absence matrix output was used to perform the Heap’s law analysis to understand the open/closedness of the pan-genome (*heaps* function of *micropan* library on R; 1000 permutations).

### SNP-based phylogenetic analyses

For the complete “global+SEAP” raw data collection, fastq reads of 1,377 *Salmonella* Paratyphi A and fasta of two RefSeq chromosomes (NC_006511 and NC_011147) were mapped against the *Salmonella* Paratyphi A AKU_12601 (FM200053.1) using Bowtie2 v2.3.5.1 [37]. Candidate SNPs were identified using SAMtools (v1.10) and BCFtools (v1.10.2) [38]. Only the homozygous, unambiguous SNPs with a Phred-quality score of >20 were selected using a customized Python script. SNPs were discarded if they had strand bias p <0.001, mapping bias p <0.001 or tail bias p <0.001 (using vcfutils.pl script, from SAMtools). SNPs located in phage or repeat regions (118.9 kb for *Salmonella* Paratyphi A AKU_12601 as described in Sajib et al. [25]) were also excluded using a customized python script. Gubbins v2.3.4 was used to detect the recombinant regions [39] and SNPs in those regions were excluded as well using the same python script, resulting in a set of 8,346 chromosomal SNPs positions for the “global+SEAP” collection (n= 1,379). All SNP alleles were extracted (fasta) using a customized python script and merged to produce SNP alignment.

Maximum likelihood trees (MLT) were built from the chromosomal SNP alignments using RAxML v8.2.12 (with the Generalized Time-Reversible model and a Gamma distribution to model site-specific rate variation; GTRGAMMA in RAxML) [15]. Support for the MLT was calculated using 100 bootstrap pseudo-analyses of the alignment. The MLT was outgroup-rooted by including the pseudo-alleles from *Salmonella* Typhi CT18 (NC_003198.1) in the alignment. Tree visualization was done using iTol v5.5 [40], including the previous Paratyphi A lineages proposed by Zhou *et al* [5].

### Bayesian analysis and identifying phylogenetically informative clades and subclades

In addition to SNP-based MLT, we investigated the population structure of the global *Salmonella* Paratyphi A collection using a Bayesian approach, implemented with the SNP alignment using fastBaps^40^. To maintain compatibility with the phylogeny, some minor modifications were made to the clustering pattern proposed by the least conservative Dirichlet prior hyperparameters on fastbaps, *optimise*.*baps*. This eventually resulted in a total of 16 different clusters. A customized python script was used to randomly select two isolates/year/cluster to represent this global collection of *Salmonella* Paratyphi A, leading to two independent sample sets of 315 isolates each. The alignment of SNP-alleles for this representative sample set was used to understand the evolutionary diverging pattern of different *Salmonella* Paratyphi A clusters over time using BEAST v1.10.4 [19]. The GTR+Γ(4) substitution model was selected for this analysis with the exponential unrelated relaxed clock as clock type and Bayesian skyline coalescent model as tree prior. The analysis considered the year of isolation as tip dates and continued for 500 million steps with sampling every 50,000 iterations. The BEAST analysis was run twice each on the two independently generated sets of isolates. The resulting log files and model parameters were analyzed on Tracer v1.7.1. TreeAnnotator v1.10 was used to generate the maximum–clade-credibility (MCC) tree [41]. The tree was visualized on FigTree v1.4.4 with a time scale. For the model with the highest posterior values (joint effective sample size (ESS) of 544) used for further analysis, time to last common ancestor (MRCA) was calculated to be 1407 AD (95% highest posterior density (HPD) interval [721.0, 1637.3]). Based on the diverging patterns suggested by the MCC tree, we assigned the clusters (defined as described above) into primary clades, secondary clades, and subclades on the MLT. However, a few visible clusters on the MLT could not be assigned to specific subclades due to a lack of clustering information from fastBaps, likely due to the low number of SNPs unique to these clusters.

### SNP-based genotyping scheme and paratype

We further divided the 16 clusters obtained from fastBAPS into 18 genotypes and identified a set of 18 SNP alleles, located in a coding sequence for conserved genes to define each assigned secondary clade and sub-clades. Each SNP allele was unique to only one subclade or, to one secondary clade and its corresponding subclades (if any). Therefore, we assigned the term “genotype” to each of the 18 secondary clades or subclades. Sorted read alignment (BAM) files generated during the SNP analysis were used to assign the genotypes for each isolate using a customized Python script, named Paratype (available at https://github.com/CHRF-Genomics/Paratype). Briefly, under BAM mode *(--mode bam*), Paratype uses *samtools index* (if bam file is not indexed), *samtools mpileup*, and *bcftools call* to extract the consensus base calls at those 18 SNP loci from the BAM file. The resulting variant call format (VCF) file is then processed to identify the presence of the defining SNP alleles and follow cladistic logic to assign the genotype of the isolate, as well as the primary clade, secondary clades, and subclade information. Paratype only considers high-quality SNP alleles (Phred score >20 and 75% read_ratio for the allele) to assign genotypes. Read_ratio is calculated by the number of high-quality alternative-allele reads on both strands, divided by the total number of high-quality reads. In addition, Paratype also has fastq mode (*--mode fastq*) where a user can provide a set of paired-end raw fastq data file (can be gzipped) and Paratype performs reference mapping (against the *Salmonella* Paratyphi AKU_12601 genome) using Bowtie and SAMtools and follows the same steps described above to detect the genotype of the isolates. Although the bam mode is the default for the tool, the fastq mode is more accurate and should be user-friendly to non-coding specializing researchers; however, it is more time-consuming. Paratype also runs on vcf mode (*--mode vcf*) which is faster, but also the least accurate if the provided SNPs are not highly trusted.

### Plasmid, resistance gene, and mutation analysis

All assembled contigs were screened with PlasmidFinder v2.1 [21] and ResFinder v3.2 [20] to detect plasmid amplicons and acquired AMR genes respectively. Both results were parsed using customized python scripts. To detect mutations in *gyrA* and *acrB* genes, we used the same Paratype script. It uses the same files used for genotyping and produces gene- and position-specific non-silent and silent mutation results.

We also explored the genomic region where the genes related to O2-antigen biosynthesis are located (860,008 – 878,865 of AKU_12601 genome). We detected all SNPs in that region with the number of isolates having those and their corresponding amino-acid changes using the Paratype. Two additional python scripts were used to count position-specific SNPs and mutations for the 18.9 kbp region.

### Data visualization and statistical analysis

R (v4.0.4) base function and several packages including dplyr, ggplot2, micropan and scatterpie were used for data visualization and statistical analysis.

## Data Availability

All data produced in the present work are contained in the manuscript

## Declarations

### Ethics approval and consent to participate

Ethical approval for the parent studies were obtained from the Bangladesh Institute of Child Health Ethical Review Committee, Nepal Health Research Council, Aga Khan University Hospital Ethics Committee and Pakistan National Ethics Committee, Stanford University Institutional Review Board, and U.S. Centers for Disease Control and Prevention. Informed written consent and clinical information were taken from adult participants and legal guardians of child participants.

### Consent for publication

Not applicable (No data from individual person was used for analysis).

### Availability of data and materials

The genome dataset supporting the conclusions of this article are available in the European Nucleotide Archive (ENA) under study accession ERP132884. The genotyping tool for *Salmonella* Paratyphi A, Paratype is available at https://github.com/CHRF-Genomics/Paratype (https://doi.org/10.5281/zenodo.5520408). Customized Python scripts and color scheme used in the manuscript are available at https://github.com/CHRF-Genomics/CHRF_Paratyphi_scripts. The metadata supporting the conclusions of this article is included in Additional file 2.

### Authors’ contributions

AMT, YH, MSIS, SKS and SS were involved in conceptualization and design of the study. MSIS performed the DNA extraction for sequencing in Bangladesh and the literature review for the global database construction. AMT, YH and MSIS performed bioinformatic analysis under supervision of SS and SKS. JRA provided continuous guidance during bioinformatic analysis. AMT and YH designed the genotyping scheme and AMT wrote the Paratype script. YH and MSIS conducted the statistical analyses and visualization. KES, JI, ZAD, SB and JRA reviewed the results. ZAD and SB reviewed genotyping scheme and the Paratype tool. AMT, YH, MSIS and SS wrote the first draft of the manuscript. KES, JI, FNQ, SPL, GD, ZAD, SB, DOG, JRA and SKS reviewed the manuscript. All authors reviewed and approved the final manuscript.

## Acknowledgements

We are thankful to Mr. Hafizur Rahman, Mr. Dipu Chandra Das and Ms. Nusrat Alam of the Child Health Research Foundation for their help with the wet-lab procedures. We are also thankful to the entire SEAP team for their unwavering support and coordination between the teams.

