## Additional file 1 for "Paratype: A genotyping framework and an open-source tool for *Salmonella* Paratyphi A"

### Supplementary Figures

Supplementary Figure 1: Pan-genome of 1379 *Salmonella* Paratyphi A genomes.

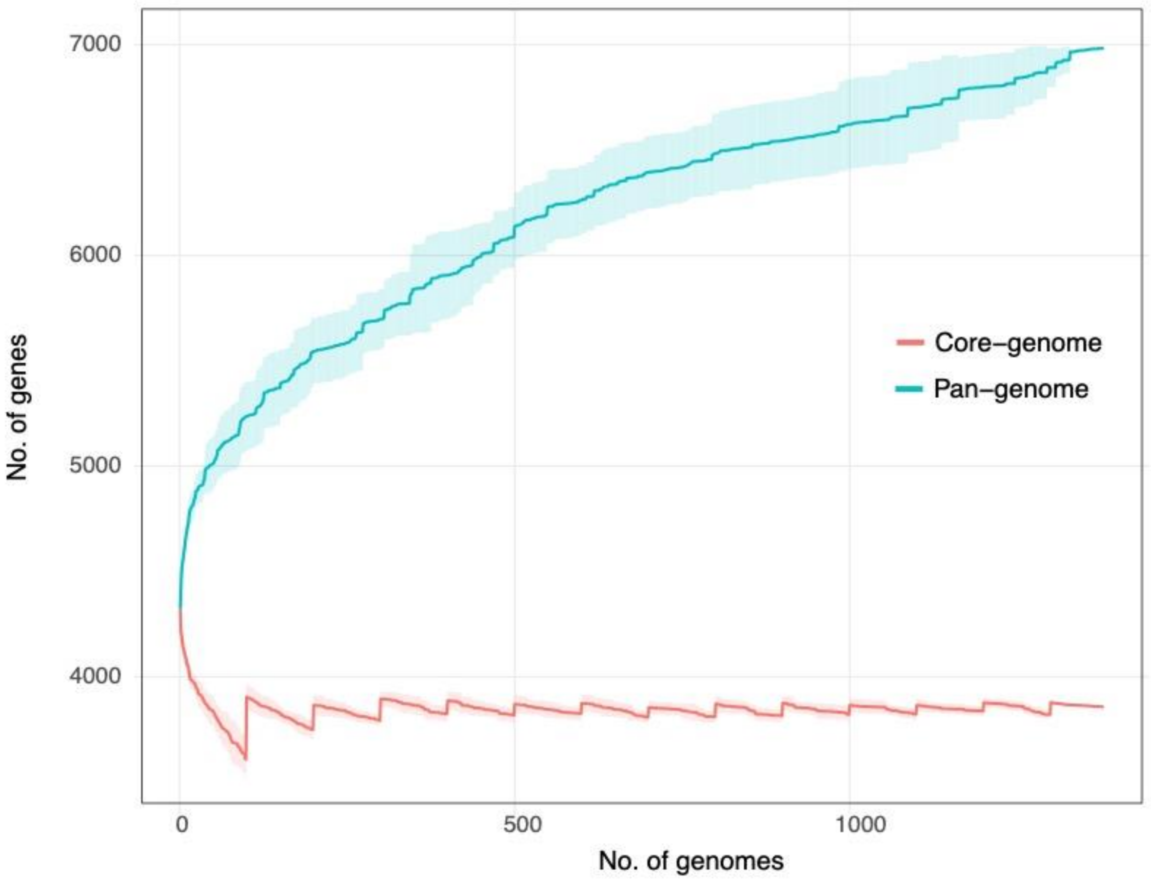


**Figure S1:** Pan-genome of 1379 *Salmonella* Paratyphi A genomes. 6983 unique genes were identified, of which 3857 were Core genes (>99% of isolates), 257 soft-core genes (95-99% of isolates), 319 shell genes (15-95% of isolates), and 2550 genes were found to be present in less than 15% of isolates.

Supplementary Figure 2: FastBAPS output for 1379 isolates used for designing the genotyping system.

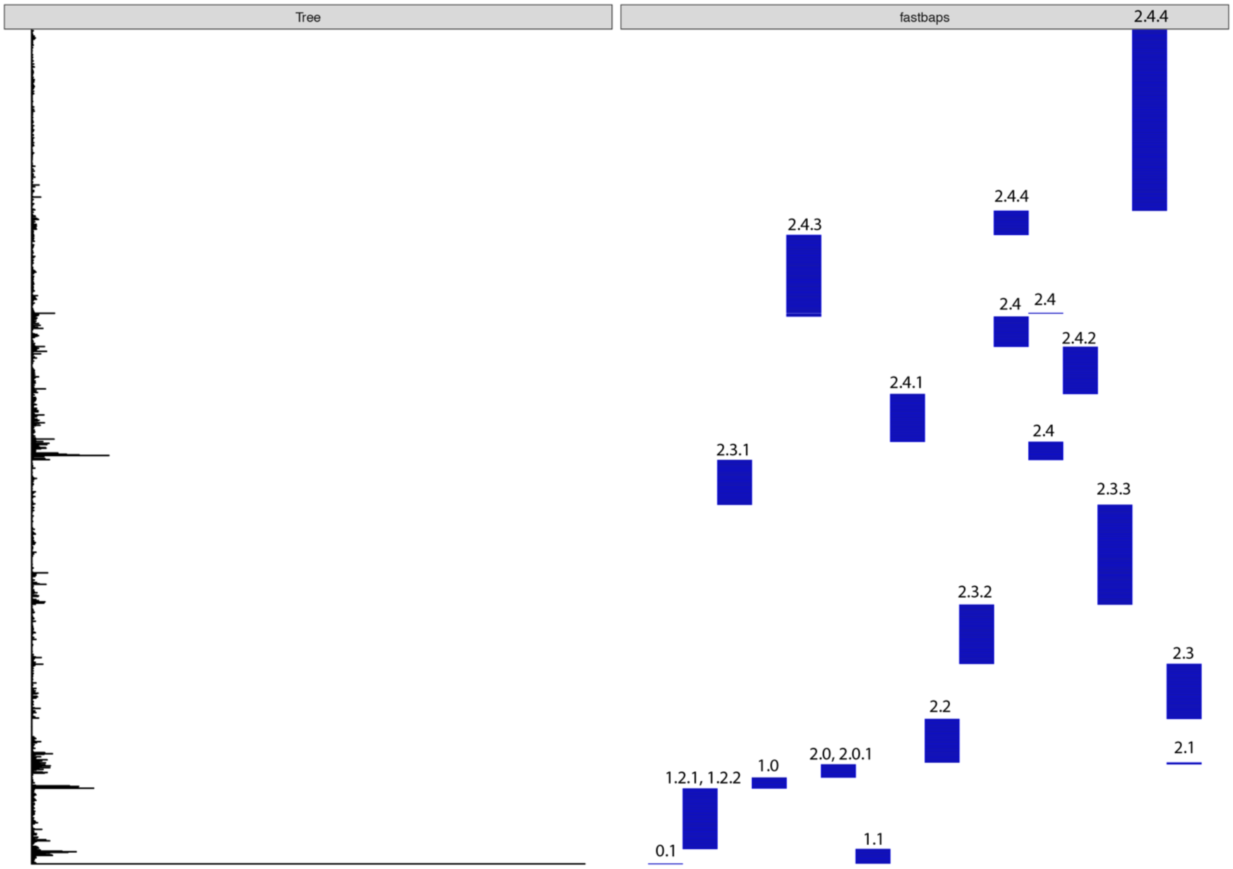


**Figure S2:** FastBAPS output for 1379 isolates used for designing the genotyping system. 16 clusters are obtained using "-optimise.baps" clustering option in fastBAPS. The plot shows the phylogenetic tree obtained from RAxML (tree) with the 16 different clusters obtained (fastbaps). The final assigned genotypes assigned to the different fastBAPS cluster are also listed.

Supplementary Figure 3: QRDR mutations among global *Salmonella* Paratyphi A isolated between 1917 and 2019.

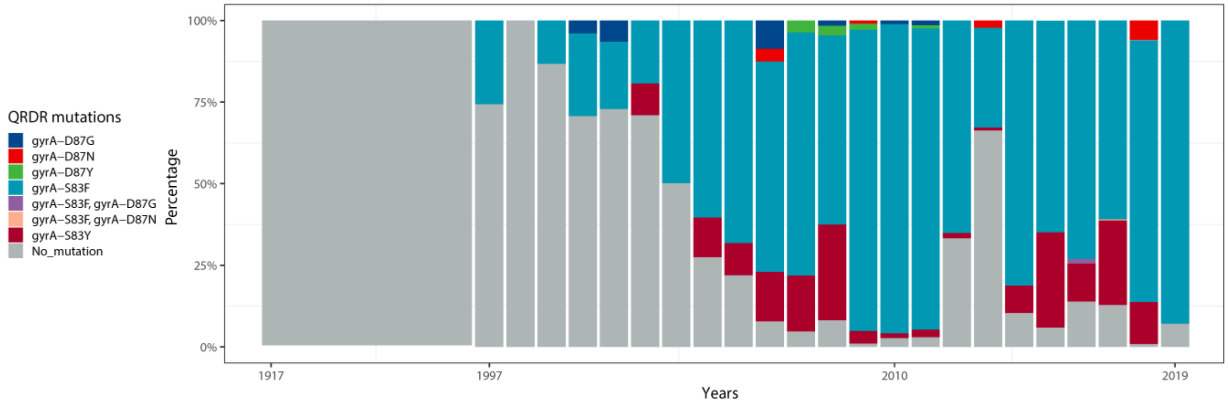


**Figure S3:** QRDR mutations among global *Salmonella* Paratyphi A isolated between 1917 and 2019. A rise in proportion of isolates with QRDR mutations has increased since mid-1990s. Since 2010, a large of majority of isolates contain atleast one QRDR mutation.

Supplementary Figure 4: Single nucleotide mutations in O2-specific region in *Salmonella* Paratyphi A.

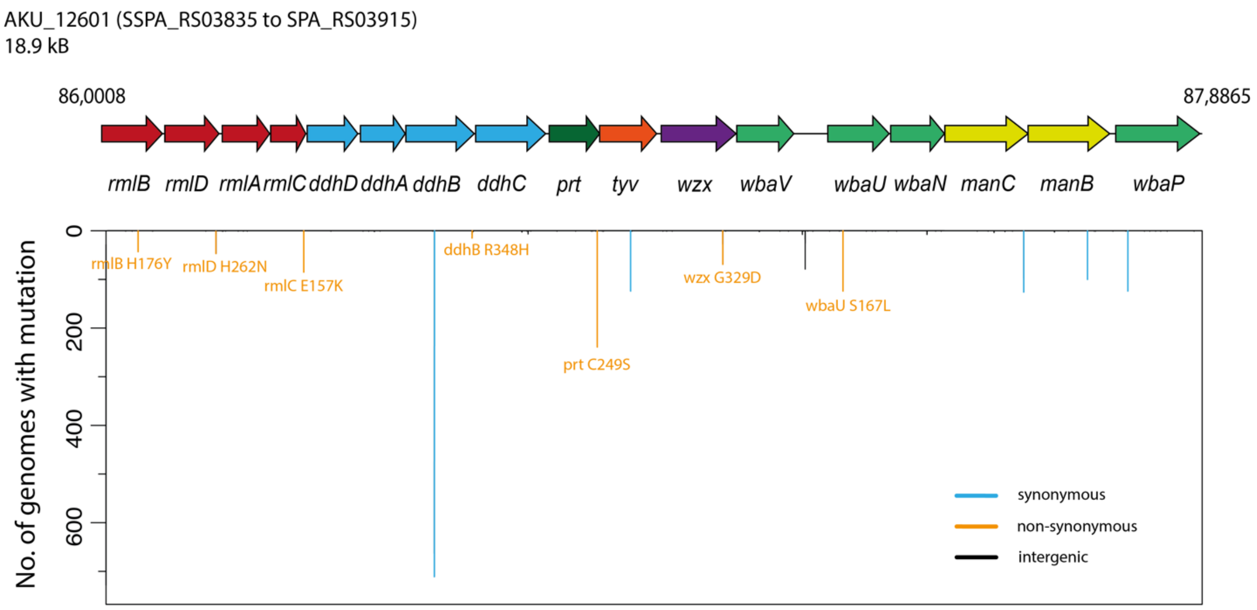


**Figure S4:** Single nucleotide mutations in O2-specific region in *Salmonella* Paratyphi A. The mutations found in the O2-specific region in *Salmonella* Paratyphi A are mapped with synonymous (cyan), non-synonymous (orange) and intergenic (black) SNPs highlighted.

### Supplementary Tables

Supplementary Table 1: Selection of isolates from the CHRF *Salmonella* Paratyphi A BioBank for whole-genome sequencing.

**Table S1:** Selection of isolates from the CHRF *Salmonella* Paratyphi A BioBank for whole-genome sequencing.

| CHRF Biobank | | | Representative sample | | |
|---|---|---|---|---|---|
| **Year** | **N** | **Freq** | **Year** | **N** | **Freq** |
| 1999 | 1 | 0% | 1999 | 1 | 0% |
| 2000 | 1 | 0% | 2000 | 1 | 0% |
| 2001 | 1 | 0% | 2001 | 1 | 0% |
| 2004 | 3 | 0% | 2004 | 2 | 1% |
| 2005 | 67 | 10% | 2005 | 44 | 13% |
| 2006 | 134 | 21% | 2006 | 42 | 12% |
| 2007 | 132 | 21% | 2007 | 55 | 16% |
| 2008 | 161 | 25% | 2008 | 67 | 19% |
| 2009 | 14 | 2% | 2009 | 13 | 4% |
| 2010 | 18 | 3% | 2010 | 17 | 5% |
| 2011 | 22 | 3% | 2011 | 22 | 6% |
| 2012 | 13 | 2% | 2012 | 13 | 4% |
| 2013 | 19 | 3% | 2013 | 19 | 5% |
| 2014 | 14 | 2% | 2014 | 13 | 4% |
| 2015 | 18 | 3% | 2015 | 17 | 5% |
| 2016 | 22 | 3% | 2016 | 21 | 6% |

Supplementary Table 2: Summary of 1,379 isolates used in the study.

**Table S2:** Summary of 1,379 isolates used in the study.

| Source | N | Freq | Country | Travel data |
|---|---|---|---|---|
| This study | 348 | 25.1 | Bangladesh | NA |
| SEAP study | 469 | 34.0 | Bangladesh, Nepal, Pakistan | NA |
| Day et al JAC 2018 [14] & Ashton et al PeerJ 2016 [8] | 254 | 18.4 | UK (Travel) | Yes (164) |
| Zhou et al PNAS 2014 [5] | 131 | 9.5 | Global | NA |
| Britto et al PLoS NTD 2018 [9] | 66 | 4.8 | Nepal | NA |
| Kuijpers et al Microb Genom 2016 [6] | 54 | 3.9 | Cambodia | NA |
| Sherchan et al ASTMH 2017 [17] | 23 | 1.7 | Nepal | NA |
| Britto et al JAC 2020 [16] | 14 | 1 | Nepal | NA |
| Yan et al. PLoS NTD 2015 [7] | 13 | 0.9 | China | NA |
| Hooda et al PLoS NTD 2019 [24] | 4 | 0.3 | Bangladesh | NA |
| Nair et al PLoS One 2020 [22] | 1 | 0.07 | UK (Travel) | Yes (1) |
| Holt et al BMC Genomics 2009 [32] | 1 | 0.07 | Pakistan | NA |
| McLelland Nature Genetics 2004 [31] | 1 | 0.07 | NA | NA |
| **Total** | **1379** | **100** | | |

Supplementary Table 3: List of alleles for the 18 genotypes.

**Table S3:** List of alleles for the 18 genotypes**.**

| Genotype | Genomic_Location | Allele | Genetic Locus | Gene name, protein |
|---|---|---|---|---|
| 0.1 | 553085 | T | SSPA_RS02400 | *aroC*, chorismite synthase |
| 1.0 | 2073394 | T | SSPA_RS10025 | cydX, cytochrome bd-I oxidase |
| 1.1 | 467038 | C | SSPA_RS01965 | *tkt*, transketolase |
| 1.2 | 1174035 | G | SSPA_RS05540 | *narG*, Nitrate reductase subunit alpha |
| 2.0 | 3387520 | G | SSPA_RS16630 | *rpsM*, 30S ribosomal protein S13 |
| 2.1 | 1576511 | G | SSPA_RS07520 | *pheT*, phenylalanine-tRNA ligase subunit beta |
| 2.2 | 2372711 | A | SSPA_RS11445 | *cyoB*, cytochrome o ubiquinol oxidase subunit I |
| 2.3 | 3386591 | T | SSPA_RS16620 | *rpsD*, 30S ribosomal subunit S4 |
| 2.4 | 865636 | T | SSPA_RS03865 | *rfbG*, CDP-glucose 4,6-dehydratase |
| 1.2.1 | 332636 | T | SSPA_RS01480 | tadA, tRNA adenosine deaminase |
| 1.2.2 | 4019735 | G | SSPA_RS19625 | *fdnG*, formate dehydrogenase-N subunit alpha |
| 2.0.1 | 2082678 | G | SSPA_RS10060 | *sucA*, 2-oxoglutarate dehydrogenase E1 component |
| 2.3.1 | 3437895 | A | SSPA_RS16940 | *trpS*, tryptophan-tRNA ligase |
| 2.3.2 | 103259 | T | SSPA_RS00455 | *rsmA*, 16S rRNA adenine demethyltransferase |
| 2.3.3 | 2419603 | G | SSPA_RS11685 | *sbcC*, exonuclease subunit |
| 2.4.1 | 1881695 | A | SSPA_RS09100 | *rpsA*, 30S ribosomal subunit S1 |
| 2.4.2 | 861876 | A | SSPA_RS03840 | *rfbD*, dTDP-4-dehydrorhamnose reductase |
| 2.4.3 | 320372 | A | SSPA_RS01410 | *lepB*, signal peptidase I |
| 2.4.4 | 1178060 | T | SSPA_RS05545 | *narH*, nitrate reductase subunit beta |

*Note: there are 19 rows for 18 genotypes, as 1.2 was divided into 1.2.1 and 1.2.2; 1.2 is not considered a unique genotype in the current version of Paratype scheme.*
